# Analysis of DNA from brain tissue on stereo-EEG electrodes reveals mosaic epilepsy-related variants

**DOI:** 10.1101/2024.07.21.24310779

**Authors:** Alissa M. D’Gama, H. Westley Phillips, Yilan Wang, Michelle Y. Chiu, Yasmine Chahine, Amanda C. Swanson, Richard S. Smith, Phillip L. Pearl, Melissa Tsuboyama, Joseph R. Madsen, Hart Lidov, Eunjung Alice Lee, Sanjay P. Prabhu, August Yue Huang, Scellig S.D. Stone, Christopher A. Walsh, Annapurna Poduri

**Affiliations:** Epilepsy Genetics Program, Department of Neurology, Boston Children’s Hospital, Boston, MA, USA; Division of Newborn Medicine, Department of Pediatrics, Boston Children’s Hospital, Boston, MA, USA; Department of Pediatrics, Harvard Medical School, Boston, MA, USA; Department of Neurosurgery, Stanford University School of Medicine, Palo Alto, CA, USA; Department of Neurosurgery, Boston Children’s Hospital, Harvard Medical School, Boston, MA, USA; Broad Institute of MIT and Harvard, Cambridge, MA, USA; Division of Genetics and Genomics, Manton Center for Orphan Disease Research, Boston Children’s Hospital, Boston, MA, USA; Program in Biological and Biomedical Sciences, Harvard Medical School, Boston, MA, USA; Division of Epilepsy and Clinical Neurophysiology, Department of Neurology, Boston Children’s Hospital, Boston, MA, USA; Department of Neurology, Harvard Medical School, Boston, MA, USA; Translational Neuroscience Center, Boston Children’s Hospital, Boston, MA, USA; Department of Pharmacology, Feinberg School of Medicine, Northwestern University, Chicago, IL, USA; Division of Neuropathology, Department of Pathology, Boston Children’s Hospital, Harvard Medical School, Boston, MA, USA; Department of Radiology, Division of Neuroradiology, Boston Children’s Hospital, Harvard Medical School, Boston, MA, USA; Howard Hughes Medical Institute, Boston Children’s Hospital, Boston, MA, USA

**Keywords:** somatic mosaicism, drug resistant epilepsy, epilepsy surgery, epilepsy genetics, deep sequencing, stereoelectroencephalography

## Abstract

Somatic mosaic variants contribute to focal epilepsy, but genetic analysis has been limited to patients with drug-resistant epilepsy (DRE) who undergo surgical resection, as the variants are mainly brain-limited. Stereoelectroencephalography (sEEG) has become part of the evaluation for many patients with focal DRE, and sEEG electrodes provide a potential source of small amounts of brain-derived DNA. We aimed to identify, validate, and assess the distribution of potentially clinically relevant mosaic variants in DNA extracted from trace brain tissue on individual sEEG electrodes.

We enrolled a prospective cohort of eleven pediatric patients with DRE who had sEEG electrodes implanted for invasive monitoring, one of whom was previously reported. We extracted unamplified DNA from the trace brain tissue on each sEEG electrode and also performed whole-genome amplification for each sample. We extracted DNA from resected brain tissue and blood/saliva samples where available. We performed deep panel and exome sequencing on a subset of samples from each case and analysis for potentially clinically relevant candidate germline and mosaic variants. We validated candidate mosaic variants using amplicon sequencing and assessed the variant allele fraction (VAF) in amplified and unamplified electrode-derived DNA and across electrodes.

We extracted DNA from >150 individual electrodes from 11 individuals and obtained higher concentrations of whole-genome amplified vs unamplified DNA. Immunohistochemistry confirmed the presence of neurons in the brain tissue on electrodes. Deep sequencing and analysis demonstrated similar depth of coverage between amplified and unamplified samples but significantly more called mosaic variants in amplified samples. In addition to the mosaic *PIK3CA* variant detected in a previously reported case from our group, we identified and validated four potentially clinically relevant mosaic variants in electrode-derived DNA in three patients who underwent laser ablation and did not have resected brain tissue samples available. The variants were detected in both amplified and unamplified electrode-derived DNA, with higher VAFs observed in DNA from electrodes in closest proximity to the electrical seizure focus in some cases.

This study demonstrates that mosaic variants can be identified and validated from DNA extracted from trace brain tissue on individual sEEG electrodes in patients with drug-resistant focal epilepsy and in both amplified and unamplified electrode-derived DNA samples. Our findings support a relationship between the extent of regional genetic abnormality and electrophysiology, and suggest that with further optimization, this minimally invasive diagnostic approach holds promise for advancing precision medicine for patients with DRE as part of the surgical evaluation.

## Introduction

Focal epilepsies, especially associated with malformations of cortical development (MCDs), are an important cause of pediatric drug-resistant epilepsy (DRE).^1^ Initial treatment with antiseizure medications (ASMs) remains largely empiric, and for individuals with focal DRE, surgical intervention provides a potential cure or palliative treatment.^2^ The pathophysiologic basis for DRE encompasses many underlying etiologies. Advances in massively parallel sequencing technologies over the past decade have accelerated progress in understanding the genetic etiologies underlying many focal epilepsies and have demonstrated the important role of somatic mosaic variants, which arise from post-zygotic mutation, in MRI-lesional and, more recently, MRI-non-lesional focal epilepsies.^3–6^

Thus far, detection of mosaic variants in focal epilepsies has relied on access to surgically resected brain tissue samples, as many of these variants are brain-limited and unable to be detected in clinically accessible tissues like blood or saliva.^7^ Given the current workflow of brain tissue-based genomics, post-operative genetic diagnosis—following an open surgical procedure for DRE—is arguably “too late” to guide precision medicine approaches and is not possible for patients who are ineligible for surgical resection or have minimally invasive interventions that do not involve brain tissue resection. Emerging diagnostic methods using alternative sources of brain-derived DNA, namely trace brain tissue from stereotactic electroencephalography (sEEG) electrodes and cell-free DNA from cerebrospinal fluid (CSF), provide an avenue for generating molecular genetic diagnosis prior to possible surgical resection and also in patients undergoing sEEG but not necessarily undergoing open cranial surgery.^8–13^ sEEG involves the temporary implantation of multiple stereotactically directed depth electrodes into the brain to help localize a patient’s seizure onset zone (SOZ). Following a period of typically several days of sEEG recording, the sEEG electrodes are explanted, at which point they have trace amounts of brain tissue attached to them. Six recent reports have demonstrated detection of potentially clinically relevant mosaic variants in DNA extracted from trace brain tissue obtained from sEEG electrodes in a total of seven patients with focal epilepsy: one patient with periventricular nodular heterotopia,^10^ one patient with multifocal non-lesional epilepsy,^9^ three patients with focal cortical dysplasia (FCD),^12,14^ one patient with a low-grade tumor,^15^ and one patient (from our group) with a diffuse frontal MCD.^16^ Here, we report the results of deep sequencing DNA from explanted sEEG electrodes from a prospectively ascertained cohort, including eleven patients undergoing invasive intracranial monitoring for DRE. In addition to identifying a mosaic *PIK3CA* variant in our previously reported case who had sEEG followed by surgical resection, we detected and validated potentially clinically relevant mosaic variants in three patients who underwent sEEG and subsequently laser interstitial thermal therapy (LITT) rather than open surgical resection and who thus did not have resected brain tissue samples available. We demonstrate the feasibility of DNA recovery and mosaic variant detection from trace brain tissue from single sEEG electrodes in multiple patients with focal DRE and highlight aspects that require optimization for application of this minimally invasive diagnostic approach to broader groups of patients.

## Materials and methods

### Patient consent

We investigated eleven consecutive pediatric patients with DRE who had sEEG electrodes implanted as part of their epilepsy surgery evaluation at Boston Children’s Hospital (BCH). One patient was previously reported.^16^ Patients or their parents provided written informed consent to enroll in the BCH Rosamund Stone Zander Translational Neuroscience Center Human Neuron Core Repository for Neurological Disorders, which is offered to all patients undergoing sEEG implantation and/or surgical resection for epilepsy at our institution. This study was approved by the BCH Institutional Review Board.

### Sample collection and DNA extraction

sEEG electrodes were implanted intracranially using standard clinical techniques and labeled using the standardized electrode nomenclature for stereoelectroencephalography applications (SENSA).^17^ Following several days of clinical monitoring for seizure and interictal spike localization, sEEG electrodes were removed through their anchor bolts (hence limiting potential contact with extracerebral tissue) and each electrode tip was cut into a 15 ml conical tube containing 10 ml cold phosphate buffered saline (PBS). Tubes containing electrodes were transported on ice and either frozen at -80^°^C until processing or immediately processed. For processing, tubes were incubated for 4 hours at 4^°^C, electrode tips were removed, and tubes were centrifuged at 3500 rpm for 10 minutes at 4^°^C. The supernatant was removed, and the cell pellet was resuspended in 10 μl PBS + 4 mM MgCl2. 3 μl was used for whole-genome amplification of DNA using primary template-directed amplification (Bioskryb Genomics) per the manufacturer’s protocol. The remaining 7 μl was used for extraction of unamplified DNA using the EZ1 DNA Tissue Kit (Qiagen) per the manufacturer’s protocol. Where available, DNA from resected brain tissue samples was extracted using the EZ1 DNA Tissue Kit, and DNA from blood or saliva samples was extracted using Qiacube (Qiagen), per the manufacturer’s protocols. DNA quantity and quality were assessed by TapeStation DNA ScreenTape analysis (Agilent).

### Immunohistochemistry

Electrodes selected for immunohistochemical analysis were collected in the operating room into 15 ml conical tubes containing ice-cold Hibernate E media (ThermoFisher, A1247601), a nutrient-rich media designed to preserve neural tissue, and transported on ice to the laboratory. Tubes containing electrodes were centrifuged at 500 rpm for 10 minutes followed by removal of 9.9 ml of the supernatant, with the remaining cell pellet resuspended in neuronal supporting tissue culture media (Neurobasal Plus + supplement media, ThermoFisher A3582901) with antibiotics Penicillin/Streptomycin (1%, ThermoFisher).

Cells were plated onto tissue culture plates pretreated with polyornithine and laminin to improve cell adherence. Cells were incubated at 37 degrees C and 5% CO2, with half media changes performed daily. On day two post-electrode collection, adherent cells were fixed with 4% paraformaldehyde in PBS for 15 minutes at room temperature, washed and stored in PBS. Immunohistochemistry was performed as previously described.^18^ Briefly, cells were blocked with goat serum and treated with Triton 0.3%, and primary antibodies were incubated overnight against NeuN (MS-RBFOX3, EMD Millipore, MAB377) and microtubule-associated protein 2 (RB-MAP2, ThermoFisher, PA517646), followed with Alexa Flour secondary antibodies. Nuclear staining was performed with Hoechst 33342 stain. Epifluorescence images were captured using the Zeiss AxioObserver Epifluorescence microscope and analysis was performed in ImageJ.

### Deep panel and exome sequencing and analysis

Deep panel sequencing was performed on a subset of samples from the eleven patients (including the one reported previously); deep exome sequencing (ES) was additionally performed on a subset of samples from eight patients (**Table 1**). Panel sequencing was performed using a custom SureSelect XT HS2 panel (Agilent) that targeted the coding exons ± 10 bp of 283 genes (target region of 852.4 kbp). The panel included genes in the mammalian target of rapamycin (mTOR) pathway, which have been associated with disease-causing mosaic variants in lesional focal epilepsies,^19^ genes in commercially available epilepsy gene panels, and genes in related cancer signaling pathways (**Supplementary Table 1**). ES was performed using a SureSelect XT HS2 Human All Exon V8 kit (Agilent). Libraries were prepared per the manufacturer’s protocols and 2 × 150 base pair paired-end sequencing was performed on Illumina HiSeq X (panel) or Illumina NovaSeq 6000 (ES) sequencers at Psomagen.

**Table 1:** Patient Cohort and Samples Used for Deep Sequencing.

| Patient/<br>Sex/Age | Notable Clinical Features | Surgical Procedure;<br>Pathology | Number<br>of<br>electrodes | Brain<br>tissue | Blood/<br>Saliva | Samples used for panel sequencing | Samples used for exome<br>sequencing |
| --- | --- | --- | --- | --- | --- | --- | --- |
| 1/F/11-15 | Right parietal choroid plexus papilloma resected at 0-5 years with subsequent encephalomalacia and gliosis, seizure onset at 6-10 years | Resection/disconnection of right parieto-occipital seizure focus and right posterior corpus callosotomy; cerebral cortex and white matter with gliosis and reactive changes | 16 | Yes | Yes | Electrodes (amplified DNA): P1b, T2A, P2dlb<br>Brian Tissue: one sample from right parietal lobe<br>Blood | Electrode (amplified DNA): P1b<br>Brain tissue: one sample from right parietal lobe |
| 2/M/16-20 | Seizure onset at 6-10 years, right frontal porencephalic cyst resected at 11-15 years with subsequent endoscopic third ventriculostomy | — | 10 | No | No | Electrodes (amplified DNA): F2d, F1aCb, F1bCc, F2aCa | Electrode (amplified DNA): F2d |
| 3/F/16-20 | DiGeorge syndrome, seizure onset at 11-15 years | Laser ablation of left posterior medial frontal seizure focus; N/A | 15 | No | Yes | Electrodes (amplified DNA): F2, F1bCb, F1cCc, F1aCa, P11b<br>Blood | Electrode (amplified DNA): F1aCa |
| 4/F/6-10 | Left temporal cavernous malformation resected at 0-5 years, seizure onset at 6-10 years | Left temporal lobectomy; cerebral cortex and white matter with gliosis, patchy neuronal dropout, and occasional maloriented and ischemic neurons, and hippocampal sclerosis | 12 | Yes | Yes | Electrodes (amplified DNA): T1a, T1b, T3aOTa, T2aA, T3bOTb, T2bH<br>Brian Tissue: samples corresponding to T3aOTa (2 samples), T1b (4 samples), T2bH (2 samples), T1a (2 samples), T2aA (3 samples), T3bOTb (2 samples) | Electrode (amplified DNA): T2bH<br>Brain tissue: sample corresponding to T2bH |
| 5/M/6-10 | Seizure onset at 0-5 years, suspected left temporal/insular FCD | Laser ablation of left temporal operculo-insular seizure focus; N/A | 16 | No | Yes | Electrodes (amplified DNA): P2b, T1d, P2alc, T1c, P2d, T1bA, P2eC, P11b<br>Blood | Electrode (amplified DNA): T1c |
| 6/M/6-10 | Seizure onset at 0-5 years, laser ablation of right subependymal heterotopia at 6-10 years, right hemiatrophy and suspected dysplasia, concern for atypical Rasmussen encephalitis | Laser ablation of right paracentral lobule and posterior insular seizure foci and needle biopsy of right superior parietal lobule; patchy neuronal loss and reactive astrogliosis | 26 | No | No | Electrodes (amplified DNA): F2a, P2ald, F1cCd, F3alaOFb | Electrode (amplified DNA): F1cCd |
| 7/M/6-10 | Tuberous Sclerosis Complex, seizure onset at 0-5 years, large left frontal tuber conglomerate resected at 0-5 years | Resection of right temporal lobe and left frontal lobe tubers/seizure foci; cortical tubers | 24 | Yes | Yes | Electrodes (amplified DNA): LF3a, LT2, RT2, LT1alb, RT1blc, LT1b, LF2bCb, RF2a, LF1aCaOF, LF2aOFa, LF3blaOF<br>Electrode (unamplified DNA): RT1blc, LF1aCaOF<br>Brian Tissue: one sample from right temporal lobe and three samples from left frontal lobe<br>Blood | — |
| 8/M/11-15 | Acute lymphocytic leukemia s/p chemotherapy, seizure onset at 0-5 years, left mesial temporal sclerosis and calcifications | Laser ablation of left mesial temporal and parietal seizure focus; N/A | 12 | No | Yes | Electrodes (amplified DNA): T3c, T2Hb, T2aHa, T1aA<br>Blood | Electrode (amplified DNA): T3aHa |
| 9/F/6-10 | Seizure onset at 0-5 years, left frontal resection at 0-5 years with FCD 1c on pathology | Laser ablation of left frontal seizure focus; N/A | 15 | No | Yes | Electrodes (amplified DNA): LF2e, LF1aOFa, LF2bCa, LF3blbOFd, RF1aCaOF<br>Blood | — |
| 10/M/16-20 | Repaired transposition of the great arteries, seizure onset at 6-10 years, encephalomalacia | Laser ablation of left fronto-parietal seizure focus; N/A | 9 | No | Yes | Electrodes (amplified DNA): F3a, F3bl, P2a<br>Blood | Electrode (amplified DNA): F3bl |

F: female, FCD: focal cortical dysplasia, M: male, N/A: not applicable

For the panel sequencing analysis, variant calling was performed using multiple methods. First, germline and mosaic variant calling was performed using SureCall (Agilent), which uses BWA for alignment to the reference human genome (hg19) and a proprietary variant caller (SNPPET) for variant calling. To achieve better sensitivity for mosaic variant detection, we further developed a customized pipeline to call mosaic single nucleotide variants (SNVs) and small insertions and deletions (indels) by MosaicHunter (version 0.1.4)^20^ and Pisces (version 5.3)^21^, respectively. For each sample, the raw fastq reads of each sample were trimmed with the AGeNT (version 2.0.5) Trimmer function before aligning to the reference genome (hg19) by BWA-MEM (version 0.7.15).^22^ The PCR duplicates were then removed with the AGeNT (version 2.0.5) LocatIT function and local indels were realigned with GATK.^23^ For mosaic variants detected by MosaicHunter and Pisces in each patient (VAF 1-30%, supported by more than three reads), we performed sample-based filtering to remove potential germline contamination: all calls that appeared in blood samples were removed from the electrode samples and resected brain tissue samples; all recurrent variants among the blood samples were also removed.

For the ES analysis, raw fastq reads of each sample were aligned with BWA-MEM (version 0.7.15)^22^ to the reference genome (hg19). The resulting bam files underwent preprocessing (mark duplicates, indel realignment, and base quality recalibration) with Picard (version 1.138)^24^ and GATK (version 3.8 and 4.1.9).^23^ Sequencing depth and the evenness of sequencing was visualized using customized Python scripts and BEDTools.^25^ MosaicHunter was used to call mosaic SNVs with the recommended ES configurations.^20^ We only considered mosaic calls with VAF <30% and excluded any calls present in dbSNP (version 138) or within 5bp of germline indels in the same patient detected by GATK HaplotypeCaller (version 3.6).^23^ To ensure a fair statistical comparison of the number of mosaic SNVs per sample, we subsampled the unamplified samples to the same mean depth as the amplified samples using Picard before rerunning MosaicHunter and applying the same filtering procedures as described above. Variant discovery and validation were conducted on the variant call set without any subsampling.

Filtered variant calls were analyzed for potentially clinically relevant variants in genes associated with human disease that matched the patient phenotype. To reduce the likelihood of false positives, we required candidate mosaic variants to be present in more than one electrode sample for panel analysis as DNA from more than one electrode sample for each patient was included on the panel. Candidate mosaic variants were manually inspected using the integrated genome viewer (IGV)^26^ prior to validation.

### Amplicon sequencing validation

Candidate variants were validated using amplicon sequencing. Validation for each candidate variant was performed for multiple samples with remaining DNA from the relevant case.

Custom primers were designed for each candidate variant using Primer3 (**Supplementary Table 2**), PCR amplification was performed using Phusion Hot Start II High-Fidelity DNA Polymerase (ThermoFisher), purification was performed using AMPure XP (Agencourt), and amplicons were sequenced on Illumina platforms using Amplicon-EZ (Genewiz). Raw fastq reads were aligned to the reference genome (hg19) using BWA-MEM (version 0.7.15),^22^ and then processed for indel realignment using GATK (version 3.6).^23^ To consider a variant as validated, we required the alternate allele of interest to have greater than three times the number of reads as the other two alternate alleles. All validated variants were further manually inspected using IGV.^26^

## Results

### Recovery and sequencing of DNA from trace brain tissue on sEEG electrodes

Eleven consecutive pediatric patients with DRE who underwent sEEG electrode implantation as part of epilepsy surgery evaluation at BCH and consented were included in this study (**Table 1** and **Fig, 1A**). sEEG electrode tips were collected at the time of explantation, resected brain tissue samples were obtained for research sequencing when open resection was performed, and blood/saliva samples were obtained when possible. Immunohistochemistry confirmed the presence of neuronal (MAP2+) as well as non-neuronal (MAP2-) cells adherent to the explanted sEEG electrodes (**Fig. 1B**). We extracted unamplified DNA and performed whole-genome amplification of DNA from the brain tissue adherent to each of >150 electrodes, with higher concentration obtained from whole-genome amplified DNA (usually >50 ng/ul) compared to unamplified DNA (usually <10 ng/ul) (**Supplementary** Fig. 1). We thus primarily used amplified DNA samples for deep sequencing but included both amplified and unamplified DNA samples for validation.

**Figure 1.**
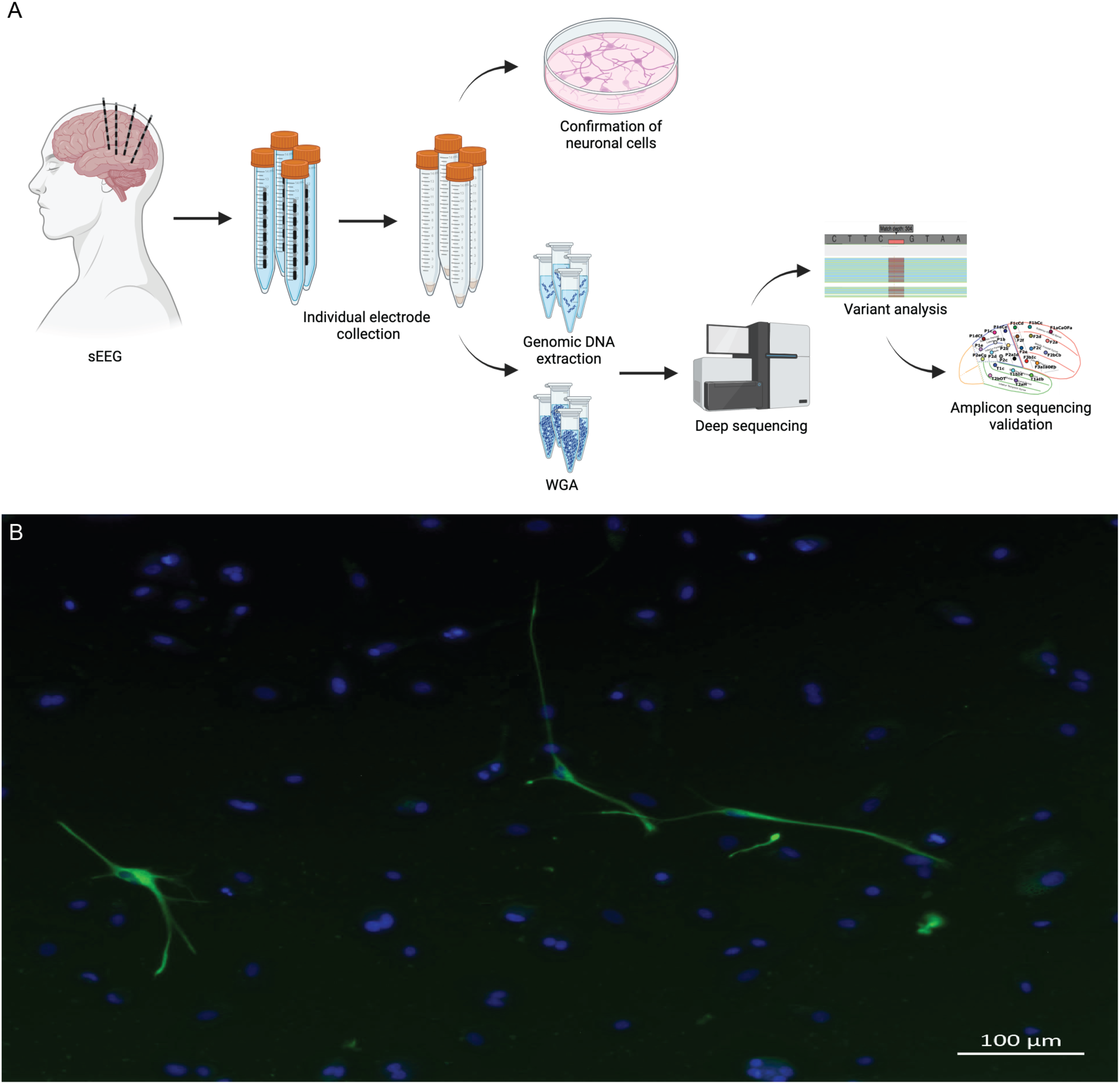
Study workflow and confirmation of neuronal cells adhered to sEEG electrodes. (A) Study workflow including sEEG procedure, individual electrode collection, confirmation of neuronal cells on electrodes, parallel extraction of genomic DNA and whole genome amplification (WGA) from individual electrodes, deep sequencing, variant analysis, and amplicon sequencing validation of variants in DNA derived from individual electrodes. Created using Biorender.com. **(B)** Epifluorescence image of sEEG electrode-derived tissue cultured cells two days post-surgical removal with antibody labeling of Microtubule-associated protein 2 (MAP2, Green), a neuron-specific cytoskeletal protein isoform for identifying neuronal cells and visualization of dendritic processes. Nuclear staining was performed with Hoechst 33342 (Blue).

We performed deep panel and/or exome sequencing using multiple samples from each patient, including at least one sample from an electrode implanted into the presumed primary seizure focus from each patient, as well as resected brain tissue and blood/saliva samples when available. Overall, the average depth of panel sequencing was 308X and of ES was 972X. For the ES samples, 63-96% of the targeted bases were sequenced to at least 500X, with the exception of the amplified sample from Patient 10, meaning that we could capture mosaic variants as low as VAF = 0.2% in these regions (**Fig. 2A**). For ES, the unamplified samples were sequenced more evenly than the amplified samples (p = 0.049, two-sided Mann-Whitney-Wilcoxon test with Bonferroni correction). Although sequencing depth did not significantly differ between amplified sEEG electrode-derived DNA samples versus unamplified samples from resected brain tissue, blood/saliva, and a few electrodes (two-sided Mann-Whitney-Wilcoxon test with Bonferroni correction, for panel sequencing, *p* = 0.76 and for ES, *p* = 0.70) (**Fig. 2B-C**), amplified samples had a significantly higher number of called mosaic SNVs and indels compared to unamplified samples (for panel sequencing, SNV p = 4.5×10^-16^, indel p = 1.1×10^-16^ and for ES, SNV p = 0.049, after subsampling the unamplified samples to the mean sequencing depth of amplified samples) (**Fig. 2D-F**). The median number of mosaic SNVs per patient identified by analysis of panel sequencing data was 223 from amplified samples versus 38 from unamplified samples, with variation among the amplified samples (SD 390.4 for amplified samples vs 3.3 for unamplified samples), suggesting that whole-genome amplification may randomly introduce artifacts that resemble mosaic variants. Thus, we required potentially clinically relevant candidate mosaic variants to be present in more than one electrode sample for panel analysis as DNA from more than one electrode sample for each patient was included on the panel. We detected and validated four potentially clinically relevant mosaic variants in three patients.

**Figure 2.**
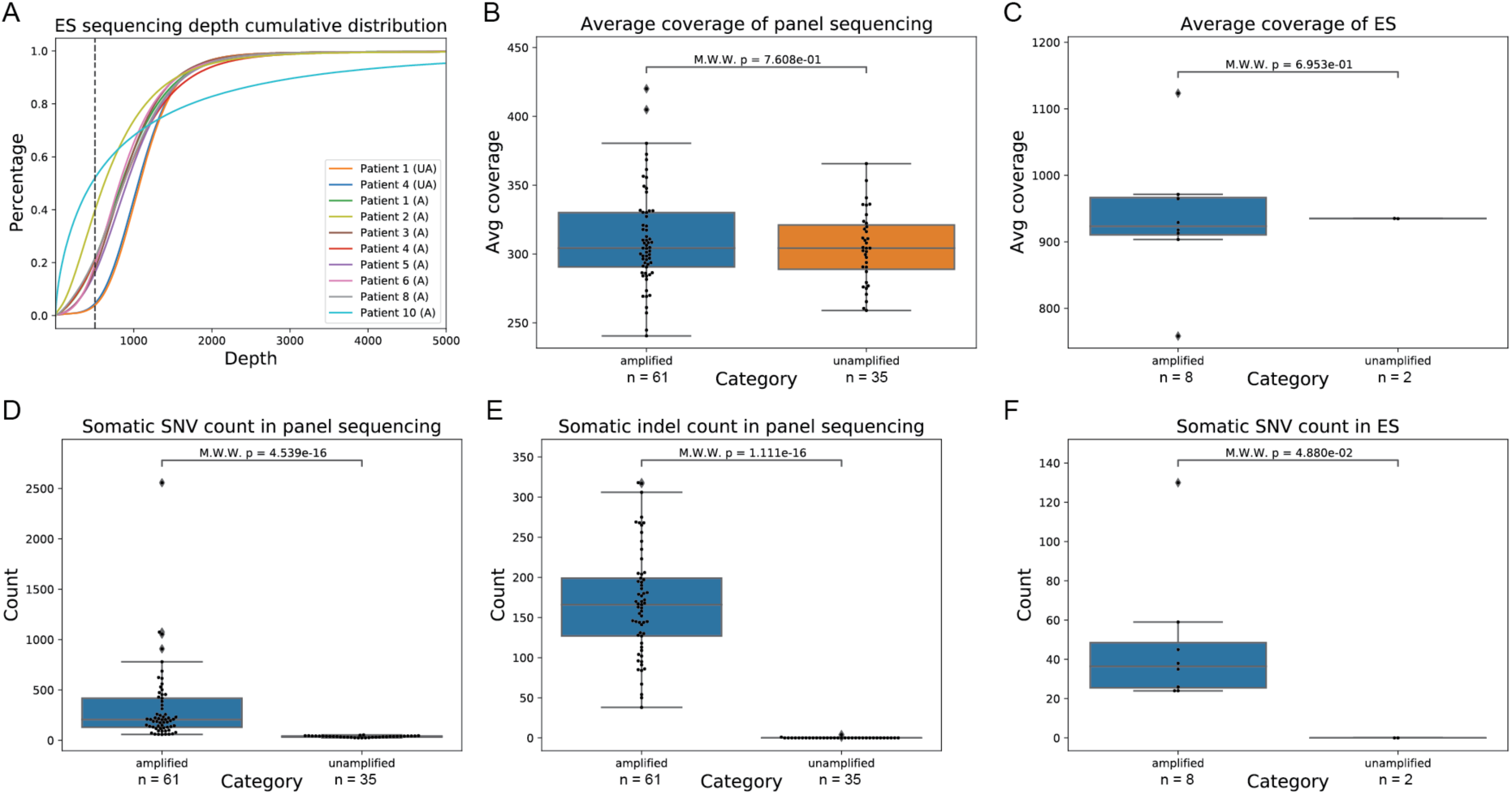
Deep sequencing coverage and variant calls. **(A)** The cumulative distribution of sequencing depth of all ES samples. A: whole-genome amplified, UA: unamplified. The dashed line indicates the percentage of targeted regions that have been sequenced with 500X or less in each sample. The depth range is restricted to up to 5000x for visualization purposes. **(B-C)** The average sequencing depth is not significantly different across amplified and unamplified samples in both panel sequencing (p = 0.76, B) and ES (p = 0.70, C). **(D-F)** The number of mosaic variants is significantly higher in amplified samples than in unamplified samples for SNVs in panel sequencing (p = 4.5×10-16, D), indels in panel sequencing (p = 1.1×10-16, E), and SNVs in ES (p = 0.049, F). M.W.W.: two-sided Mann-Whitney-Wilcoxon test with Bonferroni correction.

### Identification of clinically relevant mosaic variants from sEEG electrode-derived DNA

#### Patient 5

Patient 5 is a boy with DRE who had focal seizures with impaired awareness. MRI demonstrated blurring of the grey and white matter interface in the left posterior insula and posterosuperior temporal gyrus, suggestive of a focal cortical dysplasia (FCD) (**Fig. 3A-B**). Clinical trio ES using buccal samples had revealed a maternally inherited variant of uncertain significance (VUS) in *HUWE1* (c.10667C>T, p.P3556L), which was determined not to be clinically relevant as the HUWE1-associated intellectual disability phenotype does not closely match his clinical features and does not include FCD. He underwent epilepsy surgery evaluation, including sEEG with implantation of 16 left hemispheric electrodes (**Fig. 3C**).

**Figure 3.**
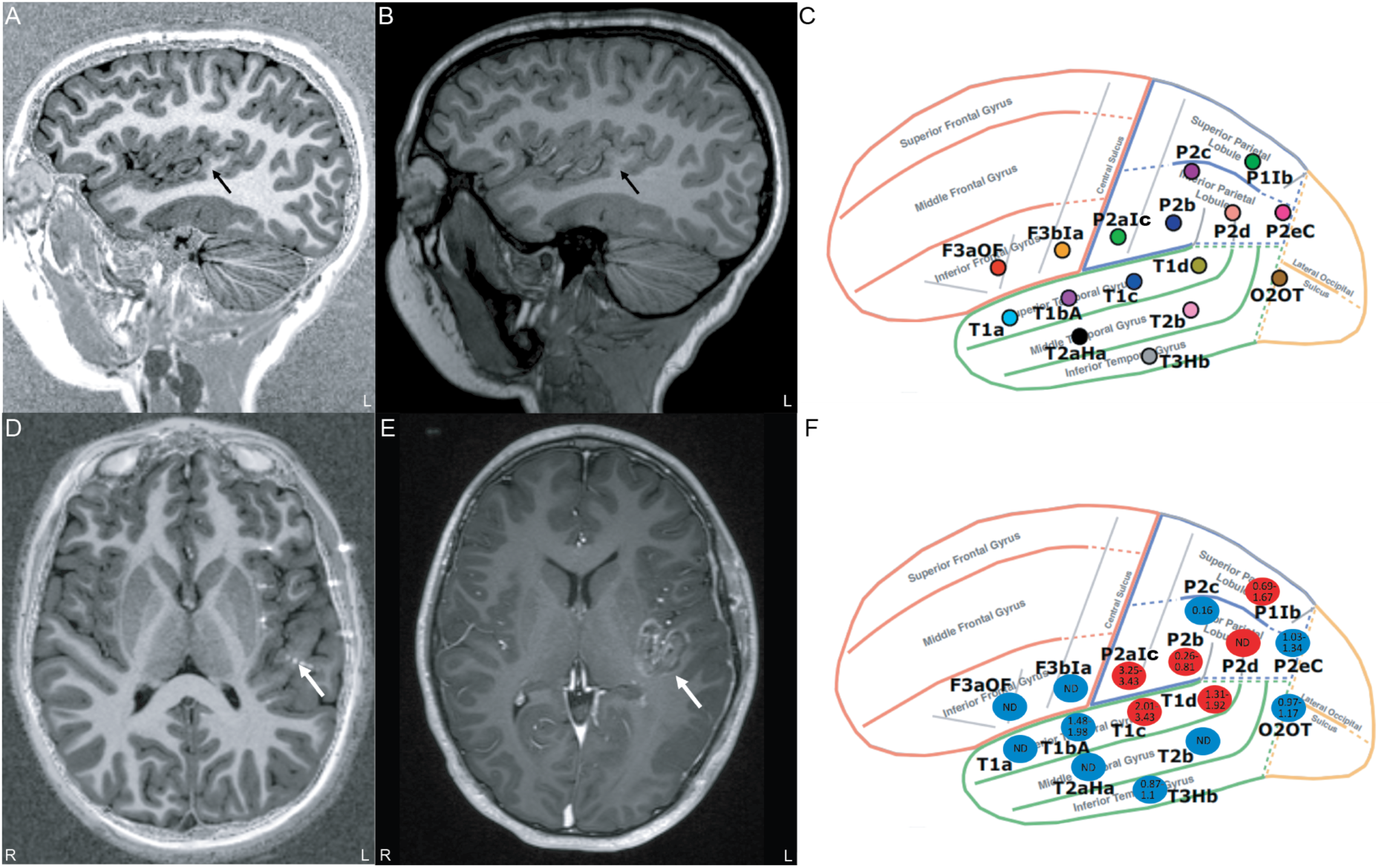
MRI, sEEG electrode placement, and *CNTNAP2* variant mosaic gradient for Patient 5. (**A-B**) Blurring of the gray and white matter interface in the left posterior insula and posterior superior temporal gyrus (arrows) on (**A)** sagittal MP2RAGE (Magnetization Prepared Rapid Acquisition Gradient Echo with two inversion times) and (**B**) conventional sagittal MPRAGE (Magnetization Prepared Rapid Acquisition Gradient Echo) images. (**C**) Lateral view of the sEEG electrode implantation plan for Patient 5 using the standardized electrode nomenclature for stereoelectroencephalography applications (SENSA) naming system. (**D**) Axial fused MP2RAGE and CT (computed tomography) image with one of the active sEEG electrodes in the suspected areas overlaid on the MP2RAGE image. (**E**) Axial post-laser interstitial thermal therapy (LITT) ablation post-contrast MPRAGE shows the treated area, which matches the abnormality seen on prior MRIs. (**F**) sEEG plan overlaid with the mosaic gradient of the detected *CNTNAP2* variant. Red ovals represent electrodes corresponding to seizure onset or spread and blue ovals represent non-involved electrodes. The number in each oval is the variant allele fraction detected in DNA extracted from that electrode. L = left, ND = not detected, R = right.

Multiple seizures were recorded, and the seizure onset zone was localized to the left temporal-opercular-insular region, in/near the location of the suspected FCD (**Fig. 3D**). After multidisciplinary discussion, the electrodes were removed and he underwent imaging-guided LITT of the seizure focus (**Fig. 3E**). The ablated tissue included areas recorded by electrodes T1c contacts 1-5, T1d contacts 1-3, P1Ib contacts 3-7, and P2d contacts 1-5. He has remained seizure-free since surgery.

For the 16 electrodes, electrode-derived amplified DNA concentrations ranged from 67.4 to 181 ng/ul and unamplified DNA concentrations ranged from 2.57 to 12.7 ng/ul. We performed deep sequencing using amplified DNA from a subset of the electrodes, including the three electrodes corresponding to the SOZ, as well as unamplified DNA from a blood sample. A mosaic missense variant in *CNTNAP2* (NM_014141.6: c.3862C>T, p.R1288C) was detected in amplified DNA from 7/8 electrodes included on the panel with VAF ranging from 0.26-3.43%. This variant is rare (gnomAD^27^ allele frequency 4.4×10^-5^) and predicted deleterious (CADD^28^ score 32). Further, the variant has previously been reported in ClinVar^29^ as a VUS (5 submissions) and in the literature as likely pathogenic based on the American College of Medical Genetics and Genomics criteria^30^ in a patient with non-lesional focal epilepsy^31^; thus, we believe this variant is potentially disease-causing for our patient. We performed subsequent amplicon sequencing using both amplified and unamplified DNA from multiple electrodes to validate and assess the distribution of the variant (**Table 2**). We validated the mosaic variant in amplified DNA from 9/16 electrodes and in unamplified DNA from 6/6 electrodes, including 5/6 electrodes corresponding to the seizure onset zone and spread. The highest VAF was seen in samples from the T1c (2.01-3.43%) and P2aIc (3.25-3.43%) electrodes, corresponding to sites of the SOZ and ictal spread, respectively (**Fig. 3F**).

**Table 2.** Variant allele frequency for *CNTNAP2* variant detected in Patient 5.

| Electrode | Source | Panel VAF (%) | Amplicon VAF (%) | Source | Amplicon VAF (%) |
| --- | --- | --- | --- | --- | --- |
| T2b | Amplified DNA | - | Not detected | Unamplified DNA | - |
| P2b<sup>b</sup> | Amplified DNA | 0.26 | 0.81 | Unamplified DNA | - |
| T1d<sup>a</sup> | Amplified DNA | 1.36 | 1.92 | Unamplified DNA | 1.31 |
| F3b1a | Amplified DNA | - | Not detected | Unamplified DNA | - |
| P2alc<sup>b</sup> | Amplified DNA | 3.25 | 3.43 | Unamplified DNA | - |
| T1c<sup>a</sup> | Amplified DNA | 3.43 | 2.01 | Unamplified DNA | - |
| P2c | Amplified DNA | - | 0.16 | Unamplified DNA | - |
| P2d<sup>a</sup> | Amplified DNA | Not detected | Not detected | Unamplified DNA | - |
| T2aHa | Amplified DNA | - | Not detected | Unamplified DNA | - |
| T3Hb | Amplified DNA | - | 0.87 | Unamplified DNA | 1.1 |
| T1a | Amplified DNA | - | Not detected | Unamplified DNA | - |
| O2OT | Amplified DNA | - | 1.17 | Unamplified DNA | 0.97 |
| T1bA | Amplified DNA | 1.98 | Not detected | Unamplified DNA | 1.48 |
| F3aOF | Amplified DNA | - | Not detected | Unamplified DNA | - |
| P2eC | Amplified DNA | 1.34 | 1.03 | Unamplified DNA | 1.03 |
| P11b<sup>b</sup> | Amplified DNA | 0.69 | 1.18 | Unamplified DNA | 1.67 |
| Blood | Unamplified DNA | Not detected | - | - | - |

<sup>a</sup> Corresponding to seizure onset zone<sup>b</sup> Corresponding to seizure spread
VAF: variant allele frequency

#### Patient 6

Patient 6 is a boy with a history of DRE. Multiple seizure types were described, including focal sensorimotor seizures and focal seizures with impaired awareness. He was initially diagnosed with a right temporal subependymal heterotopia, which was ablated (**Fig. 4A**), resulting in transient seizure freedom. Serial MRIs demonstrated progressive mild right cerebral hemispheric atrophy and dysplastic appearance of the right hemisphere, predominantly involving the frontal, parietal, and to a lesser extent, temporal lobes without well-defined borders (**Fig. 4B**). Clinical ES using buccal samples was non-diagnostic.

**Figure 4.**
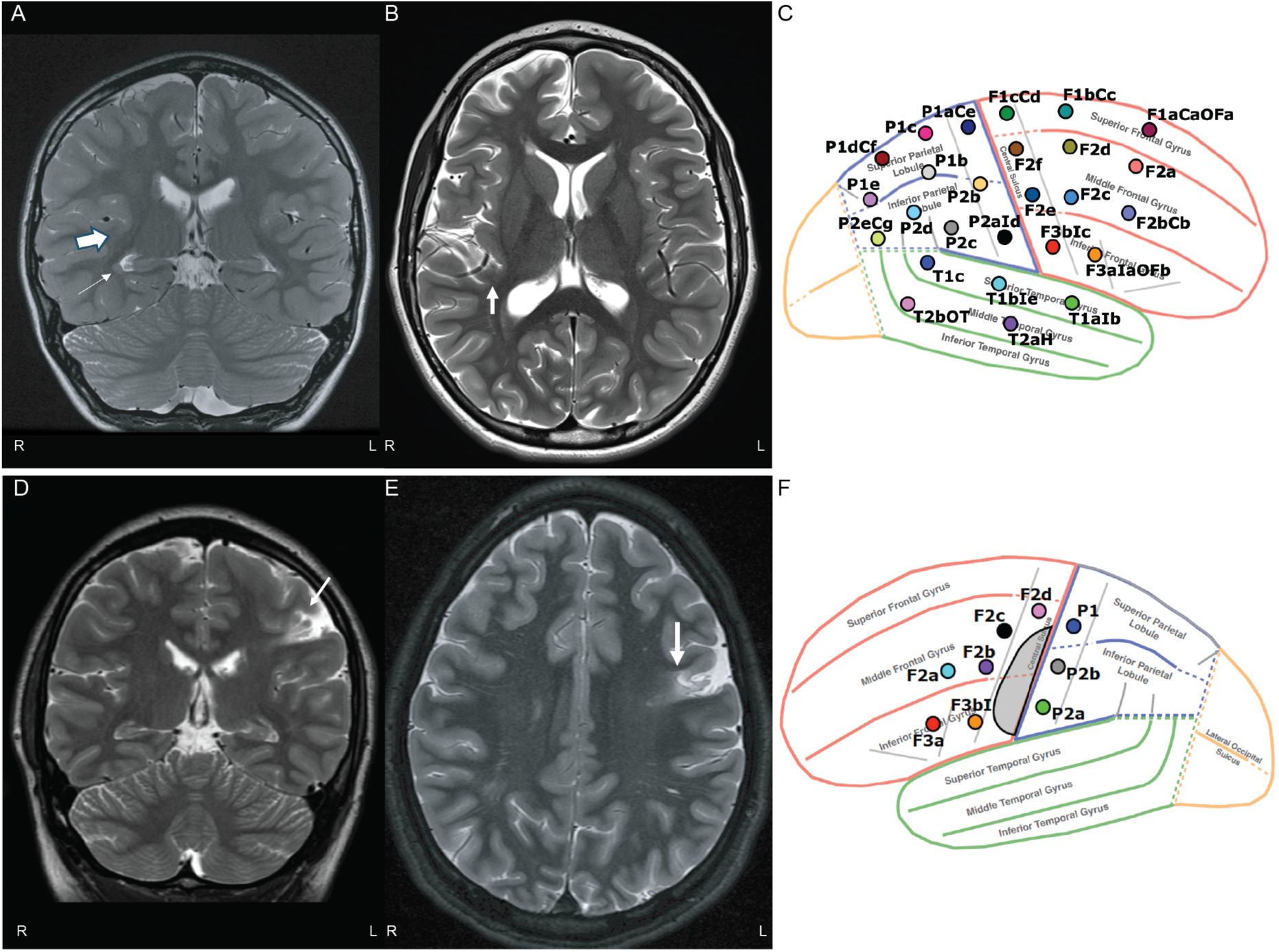
MRI and sEEG electrode placement for Patients 6 and 10. (**A**) Coronal T2-weighted MRI from an outside hospital for Patient 6 shows subependymal gray matter heterotopia adjacent to the right temporal horn (thin arrow) and subtle blurring of the gray and white matter interface in the right posterior insula and posterosuperior temporal gyrus (thick arrow). (**B**) Subsequent axial T2-weighted MRI obtained at our institution after the heterotopia had been ablated at the referring institution and before sEEG placement, again showing the subtle blurring at the gray and white matter interface in the right posterior insula (thick arrow) and right frontoparietal-predominant cerebral atrophy. (**C**) Lateral view of the sEEG electrode implantation plan for Patient 6 using the SENSA naming system. (**D-E**) Coronal (**D**) and axial (**E**) T2-weighted MRI for Patient 10 show encephalomalacia in the left frontal lobe. (**F**) Lateral view of the sEEG electrode implantation plan for Patient 10 using the SENSA naming system. The shaded grey area represents the area of encephalomalacia.

In the setting of recurrent seizures, he underwent another epilepsy surgery evaluation, including sEEG with implantation of 26 right hemispheric electrodes (**Fig. 4C**). Four seizure types were recorded: (I) epilepsia partialis continua (EPC) of the left foot, (II) focal motor seizures involving his left hemibody, (III) focal seizures with impaired awareness, and (IV) choking/gagging episodes. Seizure types I and II had SOZ in the right mesial paracentral lobule, while seizure type III involved the right anterior frontal/orbito-frontal lobe and seizure type IV involved the right posterior insula. After multidisciplinary discussion, the electrodes were removed, and he underwent LITT of the seizure foci, including regions recorded by electrodes F1bCc contacts 5-11, F1cCd contacts 1-5, P1aCe contacts 2-8, P1c contacts 1-8, P2aId contacts 1-4, and F3bIc contact 1. Pathology demonstrated patchy neuronal loss and displaced neurons with reactive astrogliosis. There was transient improvement in seizures following LITT, especially of EPC, but he has had recurrence of EPC as well as persistence of the other baseline seizure types.

For the 26 electrodes, amplified DNA concentrations ranged from 135 to 193 ng/ul and unamplified DNA concentrations ranged from 1.49 to 7.51 ng/ul. Deep sequencing using amplified DNA from a subset of the electrodes detected two potentially disease-contributory mosaic variants (**Table 3**). First, we detected a mosaic missense variant in *CIC* (NM_015125.5: c.1996C>T, p.R666C) in amplified DNA from 3/4 electrodes included on the panel with VAF ranging from 1.6-5.13%. This variant is rare (gnomAD allele frequency 4×10^-6^), predicted deleterious (CADD score 26.7), and has previously been reported in COSMIC^32^ (melanoma). Variants in *CIC* have been associated with an autosomal dominant intellectual disability disorder with seizures in some patients (OMIM^33^ 617600). We validated the mosaic variant in amplified DNA from 25/26 electrodes and in unamplified DNA from 10/11 electrodes, including 14/14 electrodes corresponding to sites with active ictal activity and ictal spread for the four seizure types.

**Table 3.** Variant allele frequency for *CIC* and *PTEN* variants detected in Patient 6.

| Electrode | Source | Panel VAF (%) | Amplicon VAF (%) | Source | Amplicon VAF (%) |
| --- | --- | --- | --- | --- | --- |
| ***CIC* variant** |  |  |  |  |  |
| F2a<sup>c</sup> | Amplified DNA | Not detected | Not detected | Unamplified DNA | 3.24 |
| F2d | Amplified DNA | - | 9.64 | Unamplified DNA | - |
| F2e | Amplified DNA | - | 2.96 | Unamplified DNA | 2.4 |
| F2f | Amplified DNA | - | 2.27 | Unamplified DNA | - |
| P2b | Amplified DNA | - | 3.21 | Unamplified DNA | - |
| P2d | Amplified DNA | - | 3.71 | Unamplified DNA | - |
| T1alb<sup>c</sup> | Amplified DNA | - | 1.54 | Unamplified DNA | - |
| F3blc<sup>a,e</sup> | Amplified DNA | - | 0.83 | Unamplified DNA | - |
| F2c<sup>c</sup> | Amplified DNA | - | 2.05 | Unamplified DNA | - |
| P1b | Amplified DNA | - | 3.1 | Unamplified DNA | 3.33 |
| P1e | Amplified DNA | - | 2.07 | Unamplified DNA | - |
| P2ald<sup>d</sup> | Amplified DNA | 1.6 | 0.78 | Unamplified DNA | 4.53 |
| T1c | Amplified DNA | - | 3.87 | Unamplified DNA | - |
| P1c<sup>a</sup> | Amplified DNA | - | 1.52 | Unamplified DNA | - |
| P2c | Amplified DNA | - | 3.5 | Unamplified DNA | 2.71 |
| T1ble | Amplified DNA | - | 2.78 | Unamplified DNA | 2.97 |
| F1bCc<sup>a</sup> | Amplified DNA | - | 1.55 | Unamplified DNA | - |
| F1cCd<sup>a</sup> | Amplified DNA | 5.13 | 4.68 | Unamplified DNA | Not detected |
| F2bCb<sup>c</sup> | Amplified DNA | - | 3.49 | Unamplified DNA | - |
| P1dCf<sup>c</sup> | Amplified DNA | - | 2.45 | Unamplified DNA | 3.12 |
| T2aH<sup>c</sup> | Amplified DNA | - | 5.61 | Unamplified DNA | - |
| T2bOT | Amplified DNA | - | 1.24 | Unamplified DNA | 2.49 |
| P1aCe<sup>a</sup> | Amplified DNA | - | 2.3 | Unamplified DNA | - |
| F1aCaOFa<sup>b</sup> | Amplified DNA | - | 5.65 | Unamplified DNA | - |
| F3alaOFb<sup>b</sup> | Amplified DNA | 3.02 | 2.51 | Unamplified DNA | 2.69, 2.62 |
| P2eCg | Amplified DNA | - | 3.95 | Unamplified DNA | 3.98 |
| ***PTEN* variant** |  |  |  |  |  |
| F2a<sup>c</sup> | Amplified DNA | 0.88 | - | Unamplified DNA | - |
| P2ald<sup>d</sup> | Amplified DNA | 2.6 | 0.44 | Unamplified DNA | - |
| F1cCd<sup>a</sup> | Amplified DNA | 1.26 | 0.42 | Unamplified DNA | - |
| F3alaOFb<sup>b</sup> | Amplified DNA | 1.45 | 0.44 | Unamplified DNA | 0.38 |

<sup>a</sup> Corresponding to seizure onset zone/spread for seizure types I (left foot EPC) and II (focal motor seizures involving left hemibody)
<sup>b</sup> Corresponding to seizure onset zone for seizure type III (focal seizure with impaired awareness)
<sup>c</sup> Corresponding to seizure spread for seizure type III (focal seizure with impaired awareness)
<sup>d</sup> Corresponding to seizure onset zone for seizure type IV (choking/gagging)
<sup>e</sup> Corresponding to seizure spread for seizure type IV (choking/gagging)
VAF: variant allele frequency

Second, we detected a mosaic missense variant in *PTEN* (NM_000314.8: c.333G>T, p.W111C) in amplified DNA from 4/4 electrodes included on the panel with VAF ranging from 0.88-2.6%. This variant is rare (not present in gnomAD), predicted deleterious (CADD score 31), and a variant at the same amino acid position (c.331T>C, p.W111R) is reported as pathogenic/likely pathogenic in ClinVar. PTEN is a negative regulator of the mTOR pathway, and variants in *PTEN* have been associated with a range of overgrowth disorders including some patients with seizures and MCDs.^1^ We validated the mosaic *PTEN* variant in amplified DNA from 3/3 electrodes and in unamplified DNA from 1/1 electrode, including 3/3 electrodes corresponding to the SOZs for the four seizure types.

#### Patient 10

Patient 10 is a man with DRE age. His seizures begin with a non-specific sensory aura followed by impaired awareness and motor involvement of his right arm, at times with evolution to bilateral tonic-clonic seizures. His medical history is notable for transposition of the great arteries, repaired in the neonatal period. MRI showed left posterior frontal and right parietal encephalomalacia, thought likely to represent remote embolic infarctions (**Fig. 4D-E**). Given this presentation, he received a diagnosis of acquired focal epilepsy and did not undergo clinical genetic testing. He underwent presurgical evaluation and his non-invasive data was concordant with the left-sided lesion, leading to sEEG with implantation of 9 left hemispheric electrodes (**Fig. 4F**). Multiple habitual seizures were recorded with onset in the left opercular insular region immediately deep and lateral to one of the encephalomalacia foci. After multidisciplinary discussion, the electrodes were removed and he underwent LITT of the seizure focus. The ablated tissue included areas recorded by electrodes F2a contact 1, F2b contacts 1-2, F3a contact 8, F3bI contacts 1-8, and P2a contacts 2-5. He was transiently seizure-free and then had recurrence of seizures that continued to be refractory to ASM treatment. He underwent an additional laser ablation to slightly expand the ablation volume, which included areas recorded by electrodes F2a, F2b, F3a, and F3bI. He had transient improvement in seizure frequency and then again had recurrence of seizures, with plans for an additional sEEG implantation.

For the 9 electrodes from his initial sEEG implantation, amplified DNA concentration ranged from 17.1 to 95.1 ng/ul and unamplified DNA concentration ranged from 2.94 to 7.2 ng/ul.

Deep ES using amplified DNA from one of the electrodes detected a potentially disease-contributory mosaic variant in *KDM6A* (NM_001291415.2: c.3203G>A, p.G1068E in the electrode F3bI, an initial site of ictal activity during the phase II recordings, at a VAF of 1.63% (**Table 4**). This variant is rare (not present in gnomAD) and predicted deleterious (CADD score 28.5). Variants in *KDM6A* have been associated with Kabuki syndrome 2 (OMIM 300867), which is characterized by intellectual disability, seizures, congenital heart disease (cardiac tissue was not available), and dysmorphic features. The variant validated in amplified DNA from 1/5 electrodes (F3bI) at a VAF of 1.13% and did not validate in unamplified DNA from 2/2 assessed electrodes or from blood.

**Table 4.** Variant allele frequency for *KDM6A* variant detected in Patient 10.

| **Electrode** | **Source** | **Exome VAF (%)** | **Amplicon VAF (%)** | **Source** | **Amplicon VAF (%)** |
| --- | --- | --- | --- | --- | --- |
| F2a | Amplified DNA | - | Not detected | Unamplified DNA | - |
| PI | Amplified DNA | - | Not detected | Unamplified DNA | - |
| F2c | Amplified DNA | - | Not detected | Unamplified DNA | Not detected |
| F3a<sup>a</sup> | Amplified DNA | - | Not detected | Unamplified DNA | Not detected |
| F3bl<sup>a</sup> | Amplified DNA | 1.63 | 1.13 | Unamplified DNA | - |
| Blood | Unamplified DNA | - | Not detected | - | - |

<sup>a</sup> Corresponding to seizure onset zone VAF: variant allele frequency

### Detection of clinically relevant germline variants

We detected previously identified germline variants in two patients. Patient 7 has DRE in the setting of Tuberous Sclerosis Complex (TSC). We detected a clinically known germline *TSC2* frameshift variant in all sequenced samples. Though TSC lesions are thought to occur due to the presence of local, tissue-specific mosaic second variants (so-called “second hits”), we did not identify a mosaic variant in the amplified DNA from electrodes, unamplified DNA from electrodes, or unamplified DNA from resected brain tissue (tuber) samples.

Patient 9 has DRE in the setting of FCD Ic demonstrated on pathology from a previous resection. We detected a clinically known germline *DEPDC5* missense variant in all sequenced samples, but despite reports of a similar second hit phenomenon for focal lesions in patients with germline *DEPDC5* variants,^34^ we did not identify a second mosaic variant in amplified DNA from electrodes. No resected brain tissue was available as the patient underwent ablation with LITT.

## Discussion

The past decade has seen increasing recognition of the important role of brain somatic mosaicism in focal DRE, initially in lesional and more recently in non-lesional cases, involving genes in the mTOR pathway as well as more recently recognized contributors, including *SLC35A2*.^35–37^ These discoveries have relied on access to surgically resected brain tissue samples. We do not know the contribution of somatic mosaic variants in patients with DRE who are ineligible for surgery or have less invasive surgical interventions that do not involve open craniotomy. The evolving practice of seizure localization using sEEG electrodes provides unique access to small amounts of DNA from these electrodes and the ability to correlate the presence and VAF of genetic variants from specific brain loci with the ictal and interictal findings in a given patient. In this study, we enrolled eleven pediatric patients with DRE undergoing sEEG implantation at our institution and aimed to detect mosaic variants in DNA extracted from the trace brain tissue recovered from the explanted sEEG electrodes, to assess the VAFs across electrodes placed in several brain regions, and to compare the feasibility of detection and validation of mosaic variants between amplified and unamplified electrode-derived DNA. In total, from these eleven individuals, we extracted amplified and unamplified DNA in parallel from over 150 individual electrodes and used deep sequencing approaches to discover and validate four mosaic variants in electrode-derived DNA from three patients who underwent laser ablation and did not have resected brain tissue samples available, in addition to the previously reported mosaic *PIK3CA* variant in a patient who did have resected brain tissue available.^16^

The use of explanted sEEG electrodes as a source of brain-derived DNA was first reported by Montier and colleagues in 2019,^10^ who identified a mosaic frameshift variant in *MEN1* in a patient with DRE and periventricular nodular heterotopia using whole-genome amplified DNA from pooled sEEG electrodes. The variant was detected at a relatively high VAF (16.7%) in the electrode-derived DNA, which allowed validation by Sanger sequencing. The authors note that the variant was not reported in previous exome sequencing (read depth not reported) using blood-derived DNA or detected in Sanger sequencing using blood-derived DNA. Subsequently, Ye and colleagues^9^ identified a mosaic nonsense variant in *KCNT1* in a patient with multifocal DRE, also using whole-genome amplified DNA from pooled sEEG electrodes. However, by collecting the electrodes into three regional pools, they were able to demonstrate a VAF gradient, with the highest VAF in the most epileptogenic region. Klein and colleagues^14,38^ recently reported identification and validation of a mosaic *MTOR* variant in a patient with DRE and FCD using whole-genome amplified DNA from neuronal nuclei isolated from pooled electrodes in the lesion/SOZ. The variant was not detected in neuronal nuclei from pooled electrodes outside the lesion, in astrocyte nuclei, or in saliva. In these case reports, the mosaic variants were detected and validated using only amplified DNA, as unamplified DNA was not extracted from the electrodes and resected brain tissue was not available. In addition, these groups used DNA extracted from pooled sEEG electrodes, limiting the resolution of the genetic findings. The presence of the *MEN1* mosaic variant in the first report was not thought to necessarily have a causal role for the individual’s epilepsy, whereas the presence of the *KCNT1* and *MTOR* mosaic variants in the latter reports were thought to have been contributing to the individuals’ epilepsy given the established roles of these genes in epilepsy.

Checri and colleagues^12^ reported detection of somatic mosaic variants, which were previously identified in resected brain tissue samples, in unamplified DNA from individual sEEG electrodes in patients with DRE and FCDIIa who underwent additional presurgical evaluations. In their first patient, the previously identified variant was detected in 3/9 assessed electrodes, all in the epileptogenic zone; in their second patient, the variant was not detected in any of the 12 assessed electrodes; and in their third patient, the variant was detected in 1/11 assessed electrodes, which was in the seizure propagation zone. The authors reported similarly low concentrations of unamplified DNA (0.2-3.9 ng/ul) from the 38 electrodes they extracted as we report here from the >150 electrodes we extracted. As in our prior report of a mosaic *PIK3CA* variant, the Checri *et al*. study reported detection of variants in electrode-derived DNA that had already been identified in resected brain tissue. Similarly, Gatesman and colleagues^15^ recently reported identification of a mosaic *FGFR1* variant from one sEEG electrode and a resected brain tissue sample in a patient with a low-grade epilepsy-associated tumor (validation was not reported). In contrast, we now report identification and validation of new variants in electrode-derived DNA in patients who did not have resected brain tissue available.

We demonstrate the ability to consistently recover trace brain tissue and extract DNA from individual sEEG electrodes and to identify and validate potentially clinically relevant mosaic variants in amplified and unamplified electrode-derived DNA. When analyzing deep sequencing data for mosaic variants, we observed that sequencing data from amplified DNA samples resulted in a significantly higher number of called mosaic variants compared to unamplified DNA samples, in the setting of similar depth of coverage, suggesting that whole-genome amplification may introduce artifacts and that sequencing of multiple independent samples from the same individual may help distinguish true somatic mutation from random artifact. When validating candidate mosaic variants, we observed that validation in amplified versus unamplified DNA from the same electrode is not always consistent—we show examples of detection in amplified but not unamplified DNA from the same electrode and vice versa. When we were able to detect the variant in both sources, the VAF was relatively similar. In our recent case report, we were unable to detect a disease-causing *PIK3CA* mosaic variant, which was identified in resected brain tissue-derived DNA, in unamplified electrode-derived DNA, although we were able to detect the variant in amplified electrode-derived DNA in 4/17 electrodes.^16^ Moreover, although in this study we were able to detect previously known disease-causing germline SNVs in amplified electrode-derived DNA, we anecdotally observed that the VAF varied across samples (e.g., for the germline *DEPDC5* variant, the VAF ranged from 28.6 to 85.6% in the amplified electrode-derived DNA samples and was called as mosaic, germline heterozygous, and germline homozygous depending on the sample; the VAF was 44.6% and called as germline heterozygous in unamplified blood-derived DNA). We were unable to detect a previously known disease-causing germline CNV in amplified electrode-derived DNA (Patient 3). Thus, although whole-genome amplification of electrode-derived DNA provides more DNA for downstream experiments, future optimization of techniques is needed. We believe extraction of unamplified DNA is important, particularly for patients where another source of unamplified brain-derived DNA (e.g., resected brain tissue) is not available, if electrode-derived DNA is to be used for clinical molecular genetic diagnosis and decision making.

In addition to our previously reported mosaic *PIK3CA* variant in a patient who underwent sEEG and resection, we discovered and validated a total of four potentially clinically relevant mosaic variants using electrode-derived DNA in three patients who underwent sEEG and LITT. In Patient 5 with DRE in the setting of an FCD, we were able to assess the *CNTNAP2* VAF distribution across amplified DNA from 16/16 electrodes and unamplified DNA from 6/16 electrodes. The *CNTNAP2* variant was detected in 2/3 electrodes corresponding to region of ictal onset and in 3/3 electrodes corresponding to seizure spread, with a relationship between the level of mosaicism and epileptogenic network suggested at single electrode resolution. Germline variants in *CNTNAP2* have been associated with an autosomal recessive neurodevelopmental disorder with epilepsy and cortical dysplasia as well as reported in autosomal dominant epilepsy; the same variant in a germline state has been reported as likely pathogenic in a patient with non-lesional focal epilepsy.^31^ To our knowledge, this is the first demonstration of a potentially disease-causing or at least disease-contributory mosaic variant discovered from individual sEEG electrodes as well as validated in amplified and unamplified DNA from single electrodes. In Patient 6, who had multiple seizure types and foci in the setting of a MCD, we validated two potentially clinically relevant mosaic variants in *CIC* and *PTEN*. Germline variants in *CIC* have been associated with an autosomal dominant intellectual disability disorder with seizures in some patients,^39^ and germline variants in *PTEN* have been associated with a range of phenotypes including MCDs and epilepsy.^40,41^ Thus, we believe these variants are at least disease-contributory, and these findings suggest that multiple mosaic variants may contribute to disease pathology in an individual patient. It is interesting to speculate if multiple mosaic variants with different distributions may contribute to epilepsy pathogenesis in patients with multifocal epilepsy, and future work is needed to investigate this hypothesis. In Patient 10, who had DRE in the setting of encephalomalacia and a history of repaired congenital heart disease, a fourth mosaic variant was detected in *KDM6A*. Germline variants in *KDM6A* have been associated with Kabuki syndrome 2, which is characterized by intellectual disability, seizures, congenital heart disease, and dysmorphic features. We did not have cardiac tissue available (although the variant was not detected in blood-derived DNA), and thus are cautious about the clinical relevance of this finding, although it may be disease-contributory.

Overall, our study demonstrates the potential utility of sequencing sEEG electrode-derived DNA as a minimally invasive genetic diagnostic approach for patients with unexplained epilepsy. This increasingly available albeit small-quantity source of DNA may be especially relevant for those individuals with DRE who are not eligible for or choose not to undergo surgical resection and thus do not have resected brain tissue available for mosaic variant assessment. We provide the first report of mosaic variant discovery using DNA derived from individual sEEG electrodes, compare amplified and unamplified electrode-derived DNA, and provide further evidence for a relationship between the degree of mosaicism, reflected in the VAF, and the degree of abnormal electrical network activity, specifically proximity to the SOZ and seizure propagation zone.

Our study has limitations, notably the relatively small number of patients and the heterogeneity of patient phenotypes, and requires validation in larger cohorts. Further, as previously noted,^12^ each sEEG electrode has multiple contacts and passes through multiple brain regions when being implanted and explanted; thus, the cells that are attached to even a single electrode may come from distinct brain regions with different electrophysiological activity. Further optimization of electrode design as well as DNA extraction, deep sequencing, and mosaic variant analysis techniques should continue to improve the sensitivity and specificity of this approach. Finally, given the relatively recent discovery of the role of mosaic variants in epilepsy-related brain lesions, our ability to ascribe causality to the mosaic variants we report in this study is limited and will require additional efforts in somatic variant curation.^42^ In the future, the minimally invasive approach we describe here holds promise for advancing precision diagnosis, which may one day become sensitive and reliable enough to implement rapidly such that the resulting data could be incorporated into the surgical decision-making process for patients with DRE. Additionally, by providing precision diagnoses, sEEG electrode-derived DNA and analysis for mosaic variants may guide surgical planning (“genetic margins”) and post-surgical prognosis using genetic data at single electrode resolution and may guide precision therapies for patients who do not undergo resection.

## Data availability

The data that support the findings of this study are available from the corresponding author, upon reasonable request.

## Acknowledgments

We thank the patients and their families who enrolled in this study.

## Funding

AMD was supported by NIH/NICHD T32 HD098061. HWP was supported by NIH/NINDS R25 NS079198 and by Credit Unions Kids at Heart. EAL and YW were supported by Allen Discovery Center program, a Paul G.Allen Frontiers Group advised program of the Paul G. Allen Family Foundation. R.S.S was supported by R00NS112604. AYH was supported by NIH/NIA R56 AG079857 and Alzheimer’s Association Research Fellowship AARF-22-972287. CAW was supported by NIH/NINDS 5R01NS035129, the Simons Foundation, and the Templeton Foundation. CAW is an Investigator of the Howard Hughes Medical Institute. This research was supported in part by the BCH Rosamund Stone Zander Translational Neuroscience Center Human Neuron Core Repository for Neurological Disorders, Credit Unions Kids at Heart, and the IDDRC (NIH P30 HD018655).

## Competing interests

The authors report no competing interests.

## Supplementary material

Supplementary material is available online.

## Notes

### Competing Interest Statement

The authors have declared no competing interest.

### Author Declarations

The IRB of Boston Children's Hospital gave ethical approval for this work.

## References

1. D’Gama AM, Poduri A. Precision Therapy for Epilepsy Related to Brain Malformations. Neurotherapeutics. Jul 2021;18(3):1548–1563. doi:10.1007/s13311-021-01122-6

2. Blumcke I, Spreafico R, Haaker G, et al. Histopathological Findings in Brain Tissue Obtained during Epilepsy Surgery. N Engl J Med. Oct 26 2017;377(17):1648–1656. doi:10.1056/NEJMoa1703784

3. Lopez-Rivera JA, Leu C, Macnee M, et al. The genomic landscape across 474 surgically accessible epileptogenic human brain lesions. Brain. Apr 19 2023;146(4):1342–1356. doi:10.1093/brain/awac376

4. Winawer MR, Griffin NG, Samanamud J, et al. Somatic SLC35A2 variants in the brain are associated with intractable neocortical epilepsy. Ann Neurol. Jun 2018;83(6):1133–1146. doi:10.1002/ana.25243

5. D’Gama AM, Walsh CA. Somatic mosaicism and neurodevelopmental disease. Nat Neurosci. Nov 2018;21(11):1504–1514. doi:10.1038/s41593-018-0257-3

6. Poduri A, Evrony GD, Cai X, Walsh CA. Somatic mutation, genomic variation, and neurological disease. Science. Jul 5 2013;341(6141):1237758. doi:10.1126/science.1237758

7. D’Gama AM, Poduri A. Brain somatic mosaicism in epilepsy: Bringing results back to the clinic. Neurobiol Dis. Jun 1 2023;181:106104. doi:10.1016/j.nbd.2023.106104

8. Ye Z, Chatterton Z, Pflueger J, et al. Cerebrospinal fluid liquid biopsy for detecting somatic mosaicism in brain. Brain Commun. 2021;3(1):fcaa235. doi:10.1093/braincomms/fcaa235

9. Ye Z, Bennett MF, Neal A, et al. Somatic Mosaic Mutation Gradient Detected in Trace Brain Tissue From Stereo-EEG Depth Electrodes. Neurology. Oct 3 2022;doi:10.1212/WNL.0000000000201469

10. Montier L, Haneef Z, Gavvala J, et al. A somatic mutation in MEN1 gene detected in periventricular nodular heterotopia tissue obtained from depth electrodes. Epilepsia. Oct 2019;60(10):e104–e109. doi:10.1111/epi.16328

11. Kim S, Baldassari S, Sim NS, et al. Detection of Brain Somatic Mutations in Cerebrospinal Fluid from Refractory Epilepsy Patients. Ann Neurol. Jun 2021;89(6):1248–1252. doi:10.1002/ana.26080

12. Checri R, Chipaux M, Ferrand-Sorbets S, et al. Detection of brain somatic mutations in focal cortical dysplasia during epilepsy presurgical workup. Brain Commun. 2023;5(3):fcad174. doi:10.1093/braincomms/fcad174

13. Ye Z, Bennett MF, Bahlo M, et al. Cutting edge approaches to detecting brain mosaicism associated with common focal epilepsies: implications for diagnosis and potential therapies. Expert Rev Neurother. Nov 2021;21(11):1309–1316. doi:10.1080/14737175.2021.1981288

14. Klein KM, Mascarenhas R, Merrikh D, et al. Identification of a mosaic MTOR variant in purified neuronal DNA in a patient with focal cortical dysplasia using a novel depth electrode harvesting technique. Epilepsia. Apr 8 2024;doi:10.1111/epi.17980

15. Gatesman TA, Hect JL, Phillips HW, et al. Characterization of low-grade epilepsy-associated tumor from implanted stereoelectroencephalography electrodes. Epilepsia Open. Feb 2024;9(1):409–416. doi:10.1002/epi4.12840

16. Phillips HW, D’Gama AM, Wang Y, et al. Somatic Mosaicism in PIK3CA Variant Correlates With Stereoelectroencephalography-Derived Electrophysiology. Neurol Genet. Feb 2024;10(1):e200117. doi:10.1212/NXG.0000000000200117

17. Stone S, Madsen JR, Bolton J, Pearl PL, Chavakula V, Day E. A Standardized Electrode Nomenclature for Stereoelectroencephalography Applications. J Clin Neurophysiol. Nov 1 2021;38(6):509–515. doi:10.1097/WNP.0000000000000724

18. Smith RS, Kenny CJ, Ganesh V, et al. Sodium Channel SCN3A (Na(V)1.3) Regulation of Human Cerebral Cortical Folding and Oral Motor Development. Neuron. Sep 5 2018;99(5):905–913 e7. doi:10.1016/j.neuron.2018.07.052

19. D’Gama AM, Woodworth MB, Hossain AA, et al. Somatic Mutations Activating the mTOR Pathway in Dorsal Telencephalic Progenitors Cause a Continuum of Cortical Dysplasias. Cell Rep. Dec 26 2017;21(13):3754–3766. doi:10.1016/j.celrep.2017.11.106

20. Huang AY, Zhang Z, Ye AY, et al. MosaicHunter: accurate detection of postzygotic single-nucleotide mosaicism through next-generation sequencing of unpaired, trio, and paired samples. Nucleic Acids Res. Jun 2 2017;45(10):e76. doi:10.1093/nar/gkx024

21. Dunn T, Berry G, Emig-Agius D, et al. Pisces: an accurate and versatile variant caller for somatic and germline next-generation sequencing data. Bioinformatics. May 1 2019;35(9):1579–1581. doi:10.1093/bioinformatics/bty849

22. Li H, Durbin R. Fast and accurate short read alignment with Burrows-Wheeler transform. Bioinformatics. Jul 15 2009;25(14):1754–60. doi:10.1093/bioinformatics/btp324

23. Van der Auwera GA, O’Connor BD. Genomics in the Cloud: Using Docker, GATK, and WDL in Terra (1st Edition). O’Reilly Media; 2020.

24. Picard Toolkit. Accessed August 4, 2023, https://broadinstitute.github.io/picard/

25. Quinlan AR, Hall IM. BEDTools: a flexible suite of utilities for comparing genomic features. Bioinformatics. Mar 15 2010;26(6):841–2. doi:10.1093/bioinformatics/btq033

26. Robinson JT, Thorvaldsdottir H, Winckler W, et al. Integrative genomics viewer. Nat Biotechnol. Jan 2011;29(1):24–6. doi:10.1038/nbt.1754

27. Karczewski KJ, Francioli LC, Tiao G, et al. The mutational constraint spectrum quantified from variation in 141,456 humans. Nature. May 2020;581(7809):434-443. doi:10.1038/s41586-020-2308-7

28. Kircher M, Witten DM, Jain P, O’Roak BJ, Cooper GM, Shendure J. A general framework for estimating the relative pathogenicity of human genetic variants. Nat Genet. Mar 2014;46(3):310–5. doi:10.1038/ng.2892

29. Landrum MJ, Lee JM, Riley GR, et al. ClinVar: public archive of relationships among sequence variation and human phenotype. Nucleic Acids Res. Jan 2014;42(Database issue):D980-5. doi:10.1093/nar/gkt1113

30. Richards S, Aziz N, Bale S, et al. Standards and guidelines for the interpretation of sequence variants: a joint consensus recommendation of the American College of Medical Genetics and Genomics and the Association for Molecular Pathology. Genet Med. May 2015;17(5):405–24. doi:10.1038/gim.2015.30

31. Kang KW, Kim W, Cho YW, et al. Genetic characteristics of non-familial epilepsy. PeerJ. 2019;7:e8278. doi:10.7717/peerj.8278

32. Tate JG, Bamford S, Jubb HC, et al. COSMIC: the Catalogue Of Somatic Mutations In Cancer. Nucleic Acids Res. Jan 8 2019;47(D1):D941–D947. doi:10.1093/nar/gky1015

33. Online Mendelian Inheritance in Man, OMIM®. McKusick-Nathans Institute of Genetic Medicine, Johns Hopkins University. Accessed August 3, 2023, https://omim.org/

34. Ribierre T, Deleuze C, Bacq A, et al. Second-hit mosaic mutation in mTORC1 repressor DEPDC5 causes focal cortical dysplasia-associated epilepsy. J Clin Invest. Jun 1 2018;128(6):2452–2458. doi:10.1172/JCI99384

35. Bonduelle T, Hartlieb T, Baldassari S, et al. Frequent SLC35A2 brain mosaicism in mild malformation of cortical development with oligodendroglial hyperplasia in epilepsy (MOGHE). Acta Neuropathol Commun. Jan 6 2021;9(1):3. doi:10.1186/s40478-020-01085-3

36. Kim JH, Park JH, Lee J, et al. Ultra-Low Level Somatic Mutations and Structural Variations in Focal Cortical Dysplasia Type II. Ann Neurol. Jun 2023;93(6):1082–1093. doi:10.1002/ana.26609

37. Lai D, Gade M, Yang E, et al. Somatic variants in diverse genes leads to a spectrum of focal cortical malformations. Brain. Aug 27 2022;145(8):2704–2720. doi:10.1093/brain/awac117

38. Mascarenhas R, Merrikh D, Khanbabaei M, et al. Detecting somatic variants in purified brain DNA obtained from surgically implanted depth electrodes in epilepsy. [Preprint] medRxiv. 2024;10.1101/2024.07.08.24310005

39. Lu HC, Tan Q, Rousseaux MW, et al. Disruption of the ATXN1-CIC complex causes a spectrum of neurobehavioral phenotypes in mice and humans. Nat Genet. Apr 2017;49(4):527–536. doi:10.1038/ng.3808

40. Dhawan A, Baitamouni S, Liu D, et al. Exploring the neurological features of individuals with germline PTEN variants: A multicenter study. Ann Clin Transl Neurol. Mar 19 2024;doi:10.1002/acn3.52046

41. Yehia L, Keel E, Eng C. The Clinical Spectrum of PTEN Mutations. Annu Rev Med. Jan 27 2020;71:103–116. doi:10.1146/annurev-med-052218-125823

42. Lai A, Soucy A, El Achkar CM, et al. The ClinGen Brain Malformation Variant Curation Expert Panel: Rules for somatic variants in AKT3, MTOR, PIK3CA, and PIK3R2. Genet Med. Nov 2022;24(11):2240–2248. doi:10.1016/j.gim.2022.07.020

