## Supplementary for "Analysis of DNA from brain tissue on stereo-EEG electrodes reveals mosaic epilepsy-related variants"

**Supplementary Table 1: Genes Included in Panel**

| | | | | |
|---|---|---|---|---|
| ABL1 | DEPDC5 | KANSL1 | NTRK2 | SCN1B |
| ACVR1 | DICER1 | KCNA2 | OFD1 | SCN2A |
| ADSL | DNAJC5 | KCNB1 | PACS1 | SCN8A |
| AKT1 | DNM1 | KCNC1 | PAFAH1B1 | SDHD |
| AKT3 | DOCK7 | KCNH1 | PCDH19 | SEC13 |
| ALDH5A1 | DYRK1A | KCNJ10 | PDGFRA | SEH1L |
| ALDH7A1 | EEF1A2 | KCNMA1 | PDPK1 | SETD2 |
| ALG13 | EGFR | KCNQ2 | PHGDH | SHANK3 |
| ALK | EHMT1 | KCNQ3 | PHOX2B | SHOC2 |
| ANKRD11 | EPCAM | KCNT1 | PIGA | SLC13A5 |
| APC | EPM2A | KCTD7 | PIGG | SLC19A3 |
| ARG1 | EPPK1 | KDM6A | PIGN | SLC25A22 |
| ARHGEF9 | ERBB2 | KDR | PIGO | SLC2A1 |
| ARX | ERBB4 | KIF7 | PIGT | SLC35A2 |
| ASNS | EZH2 | KIT | PIGV | SLC6A1 |
| ATM | FBXW7 | KLF4 | PIK3CA | SLC6A8 |
| ATP1A2 | FGF12 | KRAS | PIK3CG | SLC9A6 |
| ATP1A3 | FGFR1 | LGI1 | PIK3R1 | SMAD4 |
| ATP6AP2 | FGFR2 | LZTR1 | PIK3R2 | SMARCA2 |
| ATRX | FGFR3 | MAGI2 | PLCB1 | SMARCB1 |
| BCOR | FLNA | MAP2K1 | PLEKHS1 | SMC1A |
| BRAF | FLT3 | MAP2K2 | PMS2 | SMO |
| BRAT1 | FOLR1 | MAP3K1 | PNKP | SNAP25 |
| C12ORF57 | FOXG1 | MAP4K1 | PNPO | SOS1 |
| CACNA1A | FRRS1L | MBD5 | POLG | SPATA5 |
| CACNA1E | FUBP1 | MDM1 | PPM1D | SPRED1 |
| CACNA1G | GABBR2 | MDM2 | PPP2R5D | SPTAN1 |
| CASK | GABRA1 | MDM4 | PPT1 | SRC |
| CCND2 | GABRB2 | MECP2 | PRKAA1 | STAG2 |
| CDC73 | GABRB3 | MED12 | PRKAA2 | STK11 |
| CDH1 | GABRG2 | MEF2C | PRKAB1 | STRADA |
| CDK4 | GAMT | MEN1 | PRKAB2 | STX1B |
| CDK6 | GAPDH | MET | PRKAG1 | STXBP1 |
| CDKL5 | GATM | MFSD8 | PRKAG2 | SUFU |
| CDKN2A | GLDC | MIOS | PRKAG3 | SYNGAP1 |
| CDKN2B | GLI3 | MLH1 | PRKAR1A | SZT2 |
| CDKN2C | GLTSCR2 | MPL | PRNP | TBC1D24 |
| CHD2 | GNA11 | MSH2 | PRRT2 | TBC1D7 |
| CHRNA2 | GNAO1 | MSH6 | PTCH1 | TBL1XR1 |
| CHRNA4 | GNAQ | MTOR | PTEN | TCF4 |
| CHRNA7 | GNAS | MYC | PTPN11 | TERT |
| CHRNB2 | GOSR2 | MYCN | PURA | TP53 |
| CIC | GPC3 | MYL3 | QARS | TPP1 |
| CLCN4 | GRIN1 | NALCN | RAF1 | TSC1 |
| CLN3 | GRIN2A | NEXMIF | RB1 | TSC2 |
| CLN5 | GRIN2B | NF1 | RET | TSHZ2 |
| CLN6 | H3F3A | NF2 | RHEB | TUBB2A |
| CLN8 | HCN1 | NGLY1 | RIT1 | UBE3A |
| CNTNAP2 | HIST1H3B | NHLRC1 | RPL5 | VHL |
| CSF1R | HNF1A | NOTCH1 | RPSAP58 | WDR24 |
| CSTB | HNRNPU | NPM1 | RRAGA | WDR45 |
| CTNNB1 | HRAS | NPRL2 | RRAGB | WDR59 |
| CTSD | IDH1 | NPRL3 | RRAGC | WDR74 |
| CTSF | IDH2 | NR2F1 | RRAGD | WT1 |
| CUL4B | IQSEC2 | NRAS | SATB2 | WWOX |
| DCX | JAK2 | NRXN1 | SCARB2 | |
| DDX3X | JAK3 | NSD1 | SCN1A | |

**Supplementary Table 2: Primers Used for Amplicon Sequencing Validation**

| Gene | Primer Set |
|---|---|
| *CIC* | F: TGGCAAAGGCTATGGTTCCG<br>R: AGTGCCCATTTCAGTCCTGG |
| *CNTNAP2* | F: CTCCCAAGCCCTGTCTAACC<br>R: TATTCCATTGCCTGCCTCCC |
| *KDM6A* | F: GGATACAGTGCCGTAAAATGCT<br>R: TCACAATGCCAGATTTTCTTTGT |
| *PIK3CA* | F: CAGAGTAACAGACTAGCTAGAGAC<br>R: AGATCAGCCAAATTCAGTTA |
| *PTEN* | F: CCCACCACAGCTAGAACTTA<br>R: CCAGGAAGAGGAAAGGAAAA |

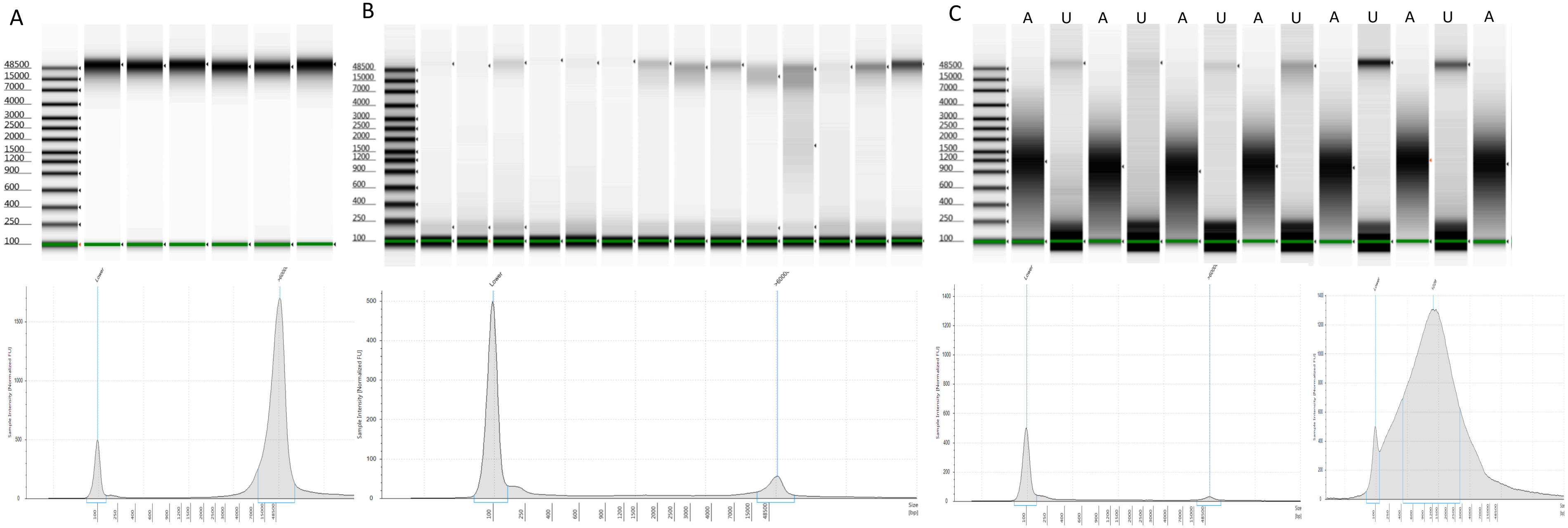


**Supplementary Figure 1**: TapeStation analysis. A) Genomic DNA (gDNA) extracted from resected brain tissue samples with well-defined >6000bp bands (top) and individual example of high amplitude >6000bp peak (bottom). B) Unamplified gDNA extracted from trace brain tissue attached to removed electrodes with less well-defined >6000bp bands (top) and individual example of lower amplitude >6000bp sample peak (bottom). C) Paired whole-genome amplified (A) and unamplified gDNA (U) from the same samples. Whole-genome amplified DNA, amplified using primary-template directed amplification from trace brain tissue attached to removed electrodes, has consistent well-defined bands centered around 1200bp (top) with individual example of high-amplitude peak centered around 1200bp (bottom) compared to the paired unamplified gDNA extractions.
